# Effectiveness of Antimicrobial Photodynamic Therapy Versus Doxycycline in the Treatment of Periodontitis on Glycaemic Control and Periodontal Parameters in Adults with Type 2 Diabetes Mellitus

**DOI:** 10.1101/2024.09.19.24313953

**Authors:** Maisie Abbott, Louise Mackenzie

## Abstract

**Background:** This meta-analysis aimed to compare the efficacy of antimicrobial photodynamic therapy (aPDT) with systemic doxycycline, as adjunctive treatments to scaling and root planing (SRP) for treating periodontitis in patients with type 2 diabetes mellitus (T2DM).

**Methods:** A search across electronic databases including PubMed, Scopus, CENTRAL, OVID EMBASE and OVID MEDLINE was carried out to identify relevant randomised controlled trials (RCTs), comparing aPDT or doxycycline combined with SRP, versus SRP alone. Data were pooled using random-effects models and indirect comparisons were retrieved using the Chi2 test for subgroup differences, to assess the impact of these treatments on glycated haemoglobin (HbA1c), probing depth (PD), and clinical attachment level (CAL).

**Results:** Eight RCTs were included. aPDT with SRP showed a significant reduction in probing depth compared to doxycycline (MD = -0.55 mm, 95% CI: -1.03 to -0.07; p = 0.02). A significant difference between the two subgroups was found (p = 0.04), suggesting aPDT was more effective. No significant differences were found for HbA1c or CAL between the two interventions.

**Conclusions:** The findings suggest that aPDT may offer greater benefits for the treatment of periodontitis in T2DM patients. This suggests that aPDT could potentially be a non-antibiotic alternative to doxycycline. However, as neither treatment influenced HbA1c, clinicians should not rely on periodontal therapy for glycaemic control.

**Layperson summary:** People with type 2 diabetes have a higher risk of getting gum disease because their bodies can’t manage blood sugar levels well. When people with type 2 diabetes get gum disease, the swelling in their gums makes it even harder to control their blood sugar levels, making their diabetes worse. Type 2 diabetics sometimes don’t respond well to regular gum treatments so dentists will also give them antibiotics to treat the gum disease. But when antibiotics are used too often, they can stop working. This is called antibiotic resistance and it’s a big problem in healthcare. Therefore, this study wanted to test if adding a special light therapy to regular gum treatments could replace using antibiotics. The results from this study found that the light therapy was better at healing parts of the gums compared to antibiotics. However, neither treatment made a big difference in blood sugar levels. This is important because it means that the light therapy could replace antibiotics for treating gum disease, reducing the problem of antibiotic resistance. However, more research is needed to see if these treatments could have any beneficial effect on blood sugar levels.

## Introduction

Periodontitis, affecting approximately 19% of the global adult population (WHO, 2023a), is characterised by a chronic and irreversible inflammatory disease state. Bacteria penetrate the supporting structures of the teeth, known as the periodontium, leading to a loss of attachment and the presence of pocketing, as shown in figure 1. Periodontitis is also a primary cause of gingival (gum) bleeding (Highfield, 2009) and tooth loss (Gasner & Schure, 2023). Research has shown that type 2 diabetes mellitus (T2DM) patients face a threefold risk of developing periodontitis (Mealey & Ocampo, 2000). T2DM is a chronic metabolic disorder, characterised by persistent hyperglycaemia due to insulin resistance with associated pancreatic beta-cell dysfunction (Goyal et al., 2023). T2DM frequently occurs in adults who have had prolonged poor glycaemic control, often because of unhealthy dietary and lifestyle choices (Sapra & Bhandari, 2023). Diabetes affects 422 million people globally, with T2DM accounting for 95% of these cases (WHO, 2023b). Research has suggested that the presence and severity of periodontitis in T2DM individuals can also lead to a reduction in glycaemic control (Preshaw et al., 2011).

**Figure 1:**
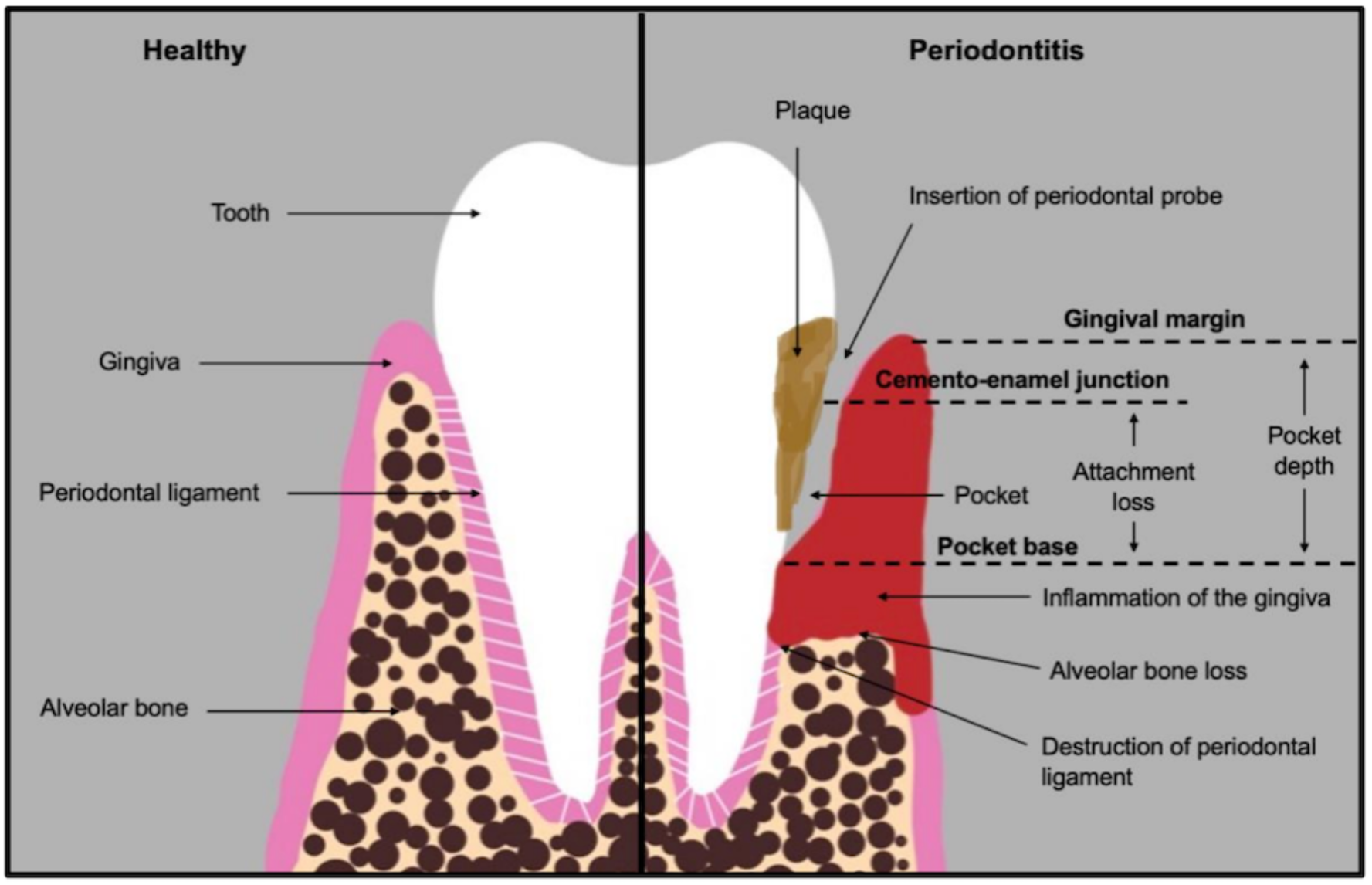
A diagram showing the comparison between a healthy periodontium and one with periodontitis, highlighting the key parameters measured in the classification of periodontitis. Based on the work of Preshaw et al., 2011, Ko et al., 2021, and Vollmer et al., 2022.

### Association between T2DM and periodontitis

Both periodontitis and T2DM share a common feature of chronic inflammation (Cecoro et al., 2020; Donath & Shoelson, 2011). Chronic hyperglycaemia in individuals with T2DM promotes the release of inflammatory markers such as TNF-a and IL-6 (Zhao et al., 2023). The release of these inflammatory markers has been shown to impair intracellular insulin signalling, and potentially contribute to insulin resistance (Hotamisligil, 2000). Systemic inflammation originating from T2DM can also compromise mucosal barriers, making periodontal tissues more susceptible to bacterial infections and subsequent inflammation (Amir et al., 2011). On the other hand, periodontitis involves localised inflammation triggered by gram-negative bacteria (Zhou et al., 2017). This leads to the overproduction of pro-inflammatory cytokines, such as IL-17, IL-6, IL-1B and TNF-a (Cochran, 2008), which can further impair glycaemic control in T2DM patients by increasing insulin resistance (Koromantzos et al., 2012).

In addition to inflammation, the oral microbiota plays a role in the association between these two conditions. Periodontitis is triggered by an imbalance in the oral microbiome, with an increase in gram-negative bacteria promoting plaque formation (Jakubovics et al., 2021). Poor glycaemic control can further exacerbate the imbalance of the oral microbiome, causing a high-glucose environment (Saeb et al., 2019), resulting in reduced oral bacterial diversity and increased levels of invasive pathogens, such as *Porphyromonas gingivalis* (Longo et al., 2018; Wang et al., 2019). Prolonged hyperglycaemia also increases glycation rates, and under oxidative stress, this can lead to the formation of advanced glycation end products (AGEs) (Ishrat et al., 2021). The AGEs, upon binding to their receptor (RAGE), accumulate in periodontal tissues (Preshaw et al., 2011), upregulating pro-inflammatory cytokine release (Lalla et al., 2001), and promoting the formation of reactive oxygen species (ROS) and matrix metalloproteinases (MMP) in the mitochondria of periodontal cells. This results in alveolar bone loss (Nonaka et al., 2017; Mei et al., 2019), as shown in figure 1.

### Treating T2DM Patients with Periodontitis

#### Scaling and Root Planing (SRP)

SRP is the gold standard treatment for individuals with periodontitis (Cobb, 1996). It is carried out by manually removing plaque and calculus from above and below the gumline, using hand instruments such as scalers and curettes (Sanz et al., 2012). However, its efficacy as a monotherapy in treating periodontitis in T2DM patients is questionable. In systemically healthy individuals, SRP has a positive effect on the healing process of periodontal tissues (Teshome & Yitayeh, 2017). However, SRP can be less effective in T2DM individuals as hyperglycaemia can lead to impaired tissue repair and healing (Geraldo et al. 2023; Lalla et al., 2000; Graves et al., 2007). Additionally, a significant limitation of SRP is its inability to effectively remove bacteria in deep periodontal pockets (Abduljabbar et al., 2017), leaving patients with a greater risk of persistent periodontal infection, heightened inflammation, and the infiltration of bacteria into the circulatory system (Luong et al., 2021).

#### Systemic Doxycycline

Doxycycline is an antibiotic that belongs to the tetracycline class (Srinath, 2015), commonly used in the treatment of periodontitis as an adjunct to SRP (BNF, 2023). It is effective against *Aggregatibacter actinomycetemcomitans* (Slots & Rams, 1990), a gram-negative bacterium (Schacher et al., 2007), which often remains in periodontal pockets even after SRP and is highly related to the aetiology and pathogenesis of periodontitis (Valle et al., 2019). It works by binding to the 30S subunit in the ribosome of the periodontopathogens, preventing the binding of aminoacyl-tRNA. By reducing the bacterial load in the periodontal pockets, doxycycline decreases the inflammatory response that leads to periodontal destruction (Srinath, 2015). In addition to its anti-microbial effects, doxycycline has also been shown to have an anti-collagenase activity. It inhibits the activity of MMPs, a type of enzyme associated with the tissue destruction process seen in periodontitis (Choi et al., 2004). By inhibiting MMPs, doxycycline helps prevent the breakdown of the periodontium (Tilakaratne & Soory, 2014), enhancing the healing process following mechanical treatments such as SRP. However, the use of antibiotics to treat periodontitis has become a subject of debate (Walters & Lai, 2015) due to concerns about the development of resistant periodontal pathogens (Jao et al., 2023), and the global health threat of antibiotic resistance (WHO, 2020).

#### Antimicrobial Photodynamic Therapy (aPDT)

To overcome the issue of antibiotic resistance, aPDT has been suggested as an alternative adjunct to SRP, with little to no risk of bacterial resistance (Theodoro et al., 2017). aPDT involves the use of three components: a photosensitiser, a light source and oxygen (Raghaendra et al., 2009). The photosensitiser, a non-toxic dye (Berakdar et al., 2012), is firstly injected into the periodontal pocket. There are many types of photosensitisers (Raghavendra et al., 2009), but for the purpose of this study methylene blue photosensitiser was the focus. When methylene blue absorbs light at a specific wavelength (665nm by a diode laser) (Raghavendra et al., 2009), it transitions to an excited triplet state, reacting with endogenous oxygen to form ROS in a controlled and targeted manner. The formation of ROS facilitates bacterial cell death (Konopka & Goslinski, 2007), leading to the elimination of periodontopathogens (Soukos et al., 2003).

### The Importance of this Meta-analysis

The presence of periodontitis in T2DM patients, significantly worsens glycaemic control (Taylor et al., 1996; Costa et al., 2017; Taboza et al., 2018) and exacerbates T2DM complications such as cardiovascular disease (Li et al., 2010). Consequently, effective management of periodontitis is crucial for improving both periodontal and systemic health outcomes in this population. While SRP is recognised as the gold standard treatment for systemically healthy individuals with periodontitis, its effectiveness in treating T2DM with periodontitis is unpredictable, emphasising the need for further research into more effective treatments. Although systemic doxycycline as an adjunct to SRP has shown to be effective in improving glycaemic control (Al-Zahrani et al., 2009; O’Connell et al., 2008), there is a risk of antibiotic resistance (Walters & Lai, 2015). aPDT has been suggested as an alternative adjunct to antibiotics, but due to the lack of scientific evidence, clinical practice guidelines do not recommend the adjunctive application of aPDT to SRP (Sanz et al., 2020). Meta-analyses and systematic reviews comparing the effects of aPDT versus doxycycline as adjuncts to SRP have been conducted, but crucially did not focus on the T2DM population (Pal et al., 2019; Akram et al., 2017). Given this gap in knowledge, this study recognised the necessity to explore the relative efficacy of aPDT vs doxycycline as adjuncts to SRP in T2DM patients. Moreover, direct head-to-head comparisons of these adjunctive interventions in randomised controlled trials (RCTs) are scarce, with only one study to date directly assessing this comparison in T2DM subjects (Ramos et al., 2015). Hence, this study employed an indirect meta-analysis approach to assess and compare the effectiveness of these two adjunctive therapies.

### Objective

To compare the effectiveness of antimicrobial photodynamic therapy (aPDT) with doxycycline as adjuncts to scaling and root planing (SRP) in individuals with T2DM for improving glycaemic control and treating periodontitis.

### Aims

1. To conduct a thorough search of electronic databases to identify relevant studies and apply a predefined inclusion criteria to select eligible studies for inclusion in the meta-analysis.
2. To extract relevant data from the included studies.
3. To perform meta-analyses using indirect comparisons, assess the overall effect sizes of adjunctive therapies, and identify heterogeneity and publication bias.
4. To interpret the findings, discuss the clinical implications and provide recommendations for future research and clinical practice.

### Hypothesis

The study hypothesised that antimicrobial photodynamic therapy (aPDT) would match or exceed the efficacy of doxycycline as adjuncts to scaling and root planing (SRP), in improving glycaemic control and periodontal parameters.

## Methods

### Eligibility Criteria

The meta-analysis aimed to address the specific question: “In adults with T2DM and periodontitis, how does aPDT and systematic administration of doxycycline each in conjunction with SRP, compare to each other in terms of improving glycemic control and periodontal health?” The eligibility criteria were structured according to the PICOS framework:

**P (Participants):** Adult human subjects (male and female) aged 18 years or older, diagnosed with T2DM and periodontitis, were included.

**I (Intervention) and C (Comparison):** Studies included in the analysis were those that provided separate comparisons of either aPDT or doxycycline, each alongside SRP, and compared to SRP alone.

**O (Outcomes):** The outcomes focused on were changes in glycated hemoglobin (HbA1c), clinical attachment level and probing depth. Reductions in all outcomes were favourable.

**S (Studies):** Only randomised controlled trials (RCTs) were considered for inclusion. Studies employing split-mouth designs were excluded, as such designs are not suitable for comparing systemic outcome measures like HbA1c, which reflect overall glycemic control rather than the localised effects that split-mouth studies are equipped to evaluate. Smokers were also excluded due to the risk factor on periodontal outcomes (Bunaes et al., 2015; Guo & Di-Pietro, 2020).

### Search Strategy

To identify studies for inclusion in the meta-analysis, a search was carried out using electronic databases including PubMed, Scopus, CENTRAL (Cochrane Central Register of Controlled Trials: Issue 3 of 12 Feb 2024), Ovid EMBASE (Excerpta Medica Database: 1974 to Feb 2024) and Ovid MEDLINE (Medical Literature Analysis and Retrieval System Online: 1946 to Feb 2024). The search strategy was developed using a combination of keywords including: ‘Type 2 Diabetes Mellitus’, ‘Periodontitis’, ‘Doxycycline’ and ‘Antimicrobial Photodynamic Therapy’, along with Boolean operators including ‘AND’ and ‘OR’. To maximise the inclusivity of the search, filters were not applied. The search results were managed using Microsoft Excel, where duplicate studies were removed before the screening process. The final search was completed on 28^th^ February 2024.

### Study Selection

A systematic approach was employed in the study selection process across three phases:

1. Title screening
2. Abstract screening
3. Full-text screening

During the first two phases, the PICOS framework was utilised to assess the relevance of each study based on its title and abstract alone. Studies that clearly addressed the population of interest, described the relevant interventions and comparisons, and stated or hinted at outcomes of relevance to the research question, were included. In cases where the relevance of a study based on its title or abstract were uncertain, the study was retained for full-text screening. During the full-text screening phase, a thorough review of the complete articles from the selected abstracts was conducted to confirm their eligibility for inclusion in the meta-analysis.

### Data Collection Process

For all studies included in the meta-analysis, outcome measures were reported as mean values and standard deviations (SD), measured at baseline and at a three-month follow-up period for the experimental and control groups. Post intervention means and SD for experimental and control groups were extracted along with their respective sample sizes. Additionally, a pre-defined data extraction form was developed using Microsoft Excel prior to collecting the data from the included studies. This form included the key characteristics of the included studies, allowing for further examination and comparison of study characteristics, ensuring consistency and similarity across the collected data.

### Assessment of Risk of Bias within Included Studies

The risk of bias was assessed by using the Cochrane Risk of Bias 2 (RoB 2) tool (22 August 2019 version) for randomised trials (Sterne et al., 2019). The following five domains were assessed: randomisation process, deviations from intended interventions, missing outcome data, measurement of outcome and selection of the reported result. Studies were considered at a low risk of bias when all criteria were met, at some concerns of bias when more than one criterion were not clearly reported, and at a high risk when more than one criterion were not met. In assessing the risk of bias across each domain and the overall study bias, both human judgement and algorithmic assessment was used.

### Summary Measures

Given that the included studies reported post-intervention outcomes using means and standard deviations on a consistent measurement scale, the data were classified as continuous. Consequently, the mean differences (MD) and relative 95% confidence intervals (CI) were calculated to quantify the effect sizes between interventions.

### Data Synthesis

Data synthesis was performed using Review Manager (RevMan [version 7.7.2] The Cochrane Collaboration, 2024). The included studies consisted of two subgroups based on the adjunctive therapy used in addition to scaling and root planing (SRP): the first subgroup consisted of studies combining antimicrobial photodynamic therapy (aPDT), in conjunction with SRP (aPDT + SRP vs SRP), whilst the second subgroup comprised studies employing systemic doxycycline alongside SRP (doxycycline + SRP vs SRP). Results from each study in both subgroups were pooled separately by using a random effects model and inverse variance to weight the subgroups in the meta-analysis (DerSimonian & Laird, 1986). Heterogeneity was evaluated using the I^2^ statistic, where values of 0% indicated no heterogeneity, and values of 25%, 50% and 75% were considered low, moderate, and high heterogeneity respectively. As only one head-to-head randomised study comparing the efficacy of aPDT and SRP with doxycycline and SRP has been published (Ramos et al., 2015), this meta-analysis relied on indirect comparisons to ascertain the relative effectiveness of these treatments, as illustrated in figure 2. Following the method described by Bucher et al. 1997, this was carried out by synthesising data from the two separate subgroups (aPDT + SRP vs SRP) and (doxycycline + SRP vs SRP), and then using the Chi^2^ test for subgroup differences, with a p value <0.05 indicating a statistically significant difference between the two subgroups.

**Figure 2:**
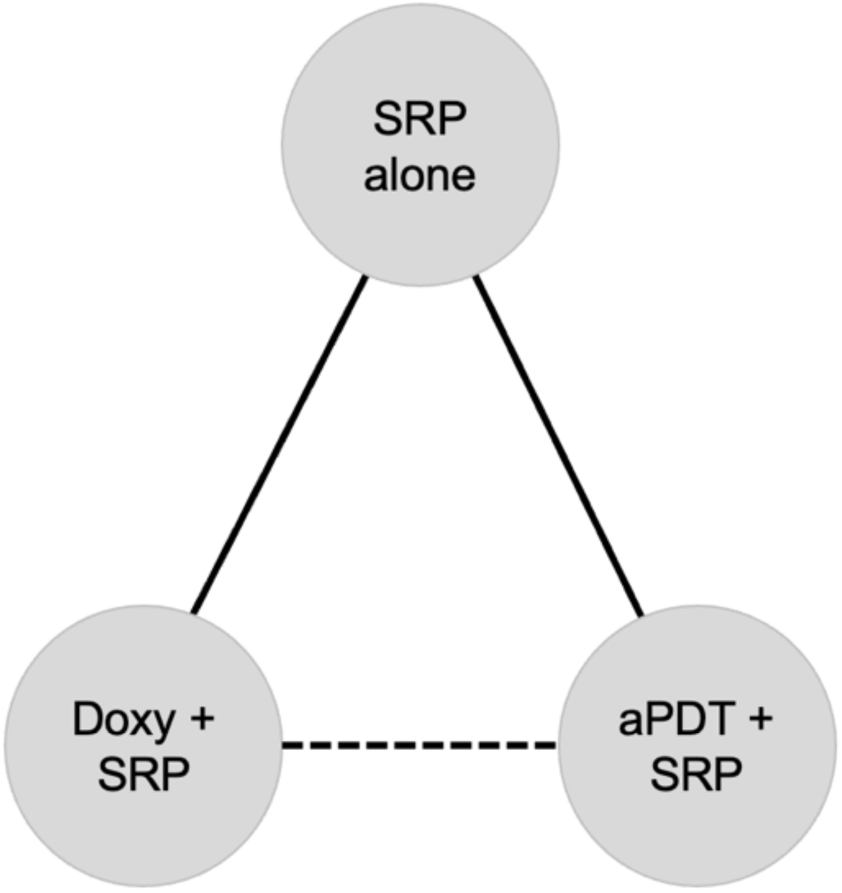
Network diagram of treatment comparisons. Each node represents a different treatment: ’SRP alone’ denotes scaling and root planing without adjunctive therapy, ‘Doxy + SRP’ represents the combination of doxycycline and scaling and root planing, and ‘aPDT + SRP’ indicates the use of antimicrobial photodynamic therapy in conjunction with scaling and root planing. Solid lines between nodes represent direct comparisons, whilst the dashed line represents the indirect comparison made between the two treatments using ‘SRP alone’ as the common comparator.

Publication bias was then assessed by visualising funnel plots containing both subgroups. If asymmetry was visualised, statistical tests were used to assess the publication bias individually for each subgroup, using the MAJOR module on Jamovi [version 2.3.28]. Egger’s regression test was used to confirm the presence of asymmetry, with a p value <0.05 indicating statistically significant asymmetry. Moreover, the fail-safe test was used to assess how many additional studies would affect the outcomes of the meta-analysis, with N corresponding to the number of studies and a p value of <0.05 indicating statistical significance. The trim and fill method was also utilised to identify any potential missing studies.

## Results

### Overview of Included Studies

#### Study Selection

Figure 3 displays the flow diagram of the study selection process. A total of 504 records were found during the literature search. After screening these records, a total of 8 studies were included in the meta-analysis.

**Figure 3:**
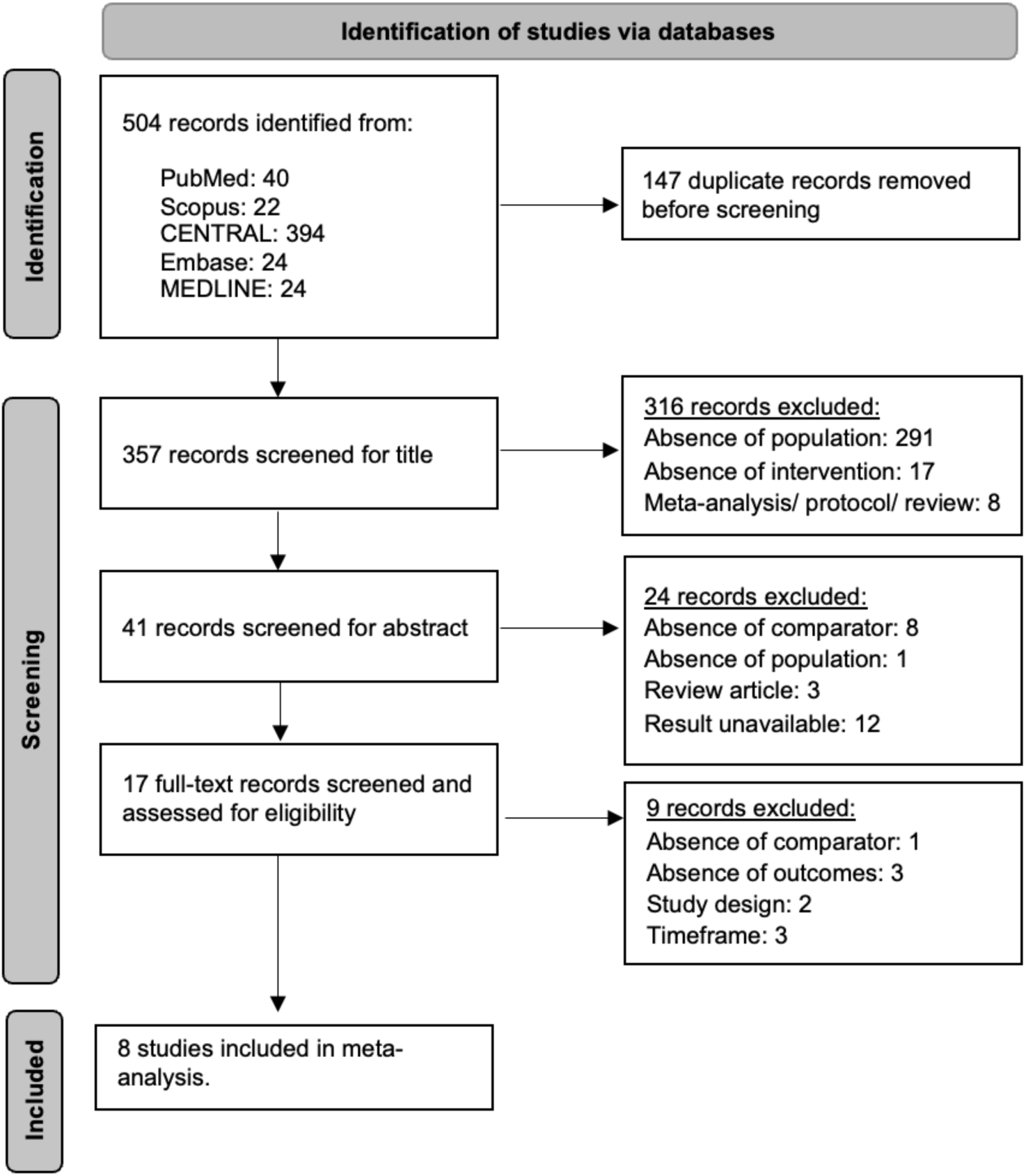
A PRISMA 2020 flow diagram (Page et al., 2021) for new systematic reviews, including searches of databases, illustrating the study selection process for the meta-analysis.

#### Network Geometry

Figure 4 shows the geometry of the network, with five studies comparing doxycycline along with SRP, compared to SRP alone (Al-Zahrani et al., 2009; Das et al., 2019; O’Connell et al., 2008; Singh et al., 2008; Tsalikis et al., 2014), and four studies comparing aPDT and SRP, compared to SRP alone (Al-Zahrani et al., 2009; Barbosa et al., 2008; Mirza et al., 2018; Thankappan et al., 2023). One study was a multi-arm study comparing both interventions to SRP alone (Al-Zahrani et al., 2009).

**Figure 4:**
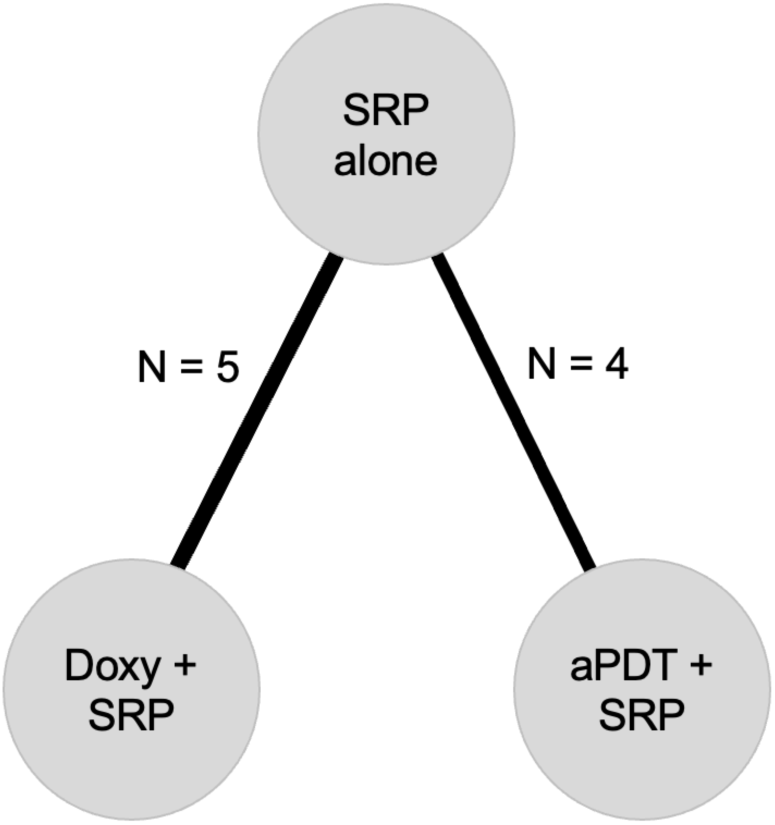
A network diagram showing the number of direct comparisons included in the meta-analysis. Each node represents an individual treatment, while the thickness of the lines connecting the nodes represents the amount of direct evidence, with its corresponding number of comparisons (N) beside it.

### Characteristics of Included Studies

#### Study Design

All studies included in the meta-analysis were RCTs. Five of the studies used computerised randomisation methods (Al-Zahrani et al., 2009, Barbosa et al., 2018, Singh et al., 2008, Thankappan et al., 2023, Tsalikis et al., 2014), two used block randomisations (Das et al., 2019, Mirza et al., 2018), and the remaining study stated that patients were randomised but did not provide details on the randomisation methods (O’Connell et al., 2008). Two studies were placebo-controlled (Barbosa et al., 2018, Tsalikis et al., 2014), whilst the remaining six were not. Moreover, six of the studies were based in a single-centre and the remaining two were multi-centre studies (Das et al., 2019, O’Connell et al., 2008). All RCTs included in the meta-analysis were parallel studies, meaning each participant only received one intervention along with SRP during the study.

#### Population

Table 1 summarises the study characteristics for all RCTs included in the meta-analysis. All studies included subjects with T2DM, with a range of baseline HbA1c values.

**Table 1:** Characteristics of Included Studies.

| Author/s | Subjects (male and female) | Exclusion criteria | Baseline HbA1c | Intervention | Control | Outcomes |
| --- | --- | --- | --- | --- | --- | --- |
| Al-Zahrani et al. 2009 | 43 (17M/26F) | Pregnant women, major diabetic complications, patients who received periodontal treatment or antibiotic therapy 6 months before study | Mean 8.80 ± 1.96 | SRP + Doxy: x2 100 mg for day 1 and then 100 mg once a day for 13 days<br>SRP + aPDT: 670nm non thermal diode with methylene blue | SRP using ultrasonic and hand instruments | Plaque and bleeding scores, PD, CAL, HbA1c |
| Barbosa et al. 2018 | 12 (33%M/67%F) | Periodontal treatment in past 6 months, prophylactic antibiotic therapy, antibiotic use within past 3 months, pregnant or breastfeeding, smokers or smoked within past 5 years, major oral cavity infections, other conditions that could affect HbA1c | Below 7% (patients taking oral hypoglycaemic drugs and insulin) | 660nm diode laser and methylene blue photosensitising agent + SRP | Ultrasound SRP and a simulating aPDT treatment lacking dye activation and laser activation | PI, PD, CAL, HbA1c and fructosamine levels |
| Das et al. 2019 | 51 (30M/21F) | Uncontrolled T2DM, periodontal therapy within the last 6 months, antibiotics within last 3 months, allergies to tetracycline, pregnant and breastfeeding women, tobacco use | Mean 7.58 ± 0.89 (SRP + doxy) and 7.58 ± 0.89 (SRP alone) | x2 100mg doxycycline initially then 100mg/day for 14 days + SRP | Oral hygiene instructions and full mouth SRP | PPD, CAL, PI, GI, FPG, PPG, HbA1c |
| Mirza et al. 2018 | 30 (20M/10F) | Pregnant or breastfeeding, antibiotic use within past 3 months, periodontal therapy within past 6 months, former/current smokers, major diabetic complications | above 6.5% | 670nm diode laser and methylene blue photosensitiser agent + SRP | SRP using ultrasonic and hand instruments along with chlorohexidine rinse | HbA1c, AGEs, PI, BoP, PD, CAL |
| O'Connell et al. 2008 | 30 (14M/16F) | Use of antibiotics or periodontal treatment within past 6 months, smoking within past 5 years, pregnant or breastfeeding, major diabetic complications, and cocomitant medical therapy | Mean 11.2% | x2 100mg doxycycline initially then 100mg/day for 14 days + SRP | SRP using ultrasonic and hand instruments | CAL, PD, BOP, PI, suppuration, missing teeth, HbA1c, FPG, serum biomarkers |
| Singh et al. 2008 | 45 (M/F NA) | Uncontrolled T2DM, periodontal treatment within past 6 months, antibiotic use within past 3 months, less than 16 teeth | Mean 8.3 ± 0.7 (SRP + doxy) and 7.9 ± 0.7 (SRP alone) | x2 100mg doxycycline initially then 100mg/day for 14 days + SRP | Oral hygiene instructions and full mouth SRP | PI, GI, PPD, CAL, FBG, PPG, Hba1c |
| Thankappan et al. 2023 | 16 (10M/6F) | Patients who received periodontal, antibiotic or aPDT in the last 90 days. Current smokers, pregnant or breast feeding women, allergy to dye, uncontrolled T2DM | 7% to 10% | HELBO blue diode laser 660nm with photosensitiser containing methylene blue | SRP using ultrasonic and hand instruments | PI, GI, PPD, CAL, HbA1c |
| Tsalikis et al. 2014 | 66 (38M/28F) | T1DM, antibiotic use within past 3 months, smokers, other systemic diseases, pregnant or breastfeeding, periodontal treatment within past 6 months | below 7.5% | x2 100mg doxycycline initially then 100mg/day for 14 days + SRP | SRP using ultrasonic and hand instruments and placebo | PPD, gingival recession, CAL, BoP, HbA1c, MMP-8, subgingival species |

Abbreviations: M, male; F, female; SRP, scaling and root planing; aPDT, antimicrobial photodynamic therapy; doxy, doxycycline; PI, plaque index; GI, gingival index; PPD, periodontal probing depth; CAL, clinical attachment level; PD, probing depth; FPG, fasting plasma glucose; PPG, postprandial glucose; BOP, bleeding on probing.

#### Interventions

In all studies where systemic doxycycline was administered as an intervention (Al-Zahrani et al., 2009, Das et al., 2019, O’Connell et al., 2008, Singh et al., 2008, Tsalikis et al., 2014), the treatment protocol consisted of an initial dose of 200mg followed by a daily dose of 100mg for a duration of 14 days. In all studies employing aPDT, methylene blue was utilised as the photosensitiser, with two studies using a 660nm diode laser (Barbosa et al., 2008, Thankappan et al., 2023) and two using a 670nm diode laser (Al-Zahrani et al., 2009, Mirza et al., 2018). In all studies, SRP was carried out using hand and/or ultrasonic instruments. One study also used a chlorohexidine rise (Mirza et al., 2018).

#### Outcomes

All the studies included in the meta-analysis measured HbA1c levels to evaluate the impact of the interventions on T2DM management and evaluated clinical attachment level and probing depth to determine improvement in periodontal health.

### Risk of Bias Assessment

The outcomes for the assessment of risk of bias are presented in figure 5. In domain 1, all studies randomised participants to their intervention groups, yet some concerns were noted in six studies due to insufficient information about whether the allocation sequence was concealed until participants were enrolled and assigned to interventions. In domain 2, four of the studies implemented double blinding, ensuring both participants and intervention administrators were unaware of the treatment assignments. In contrast, the remaining studies had unclear blinding procedures, which may have led to deviations from the intended intervention. Domain 3 showed a low risk of bias across all studies, as data for all, or nearly all randomised patients were available and accounted for within the analysis. Domain 4 also demonstrated a low risk of bias across all studies, as outcome measurements were consistently based on established clinical parameters, such as HbA1c, ensuring valid results. In domain 5, three studies were noted for their low risk of bias due to their adherence to a pre-specified protocol mentioned in their methodologies. However, the remaining studies did not provide sufficient information on their reporting plans, potentially allowing selective reporting of results.

**Figure 5:**
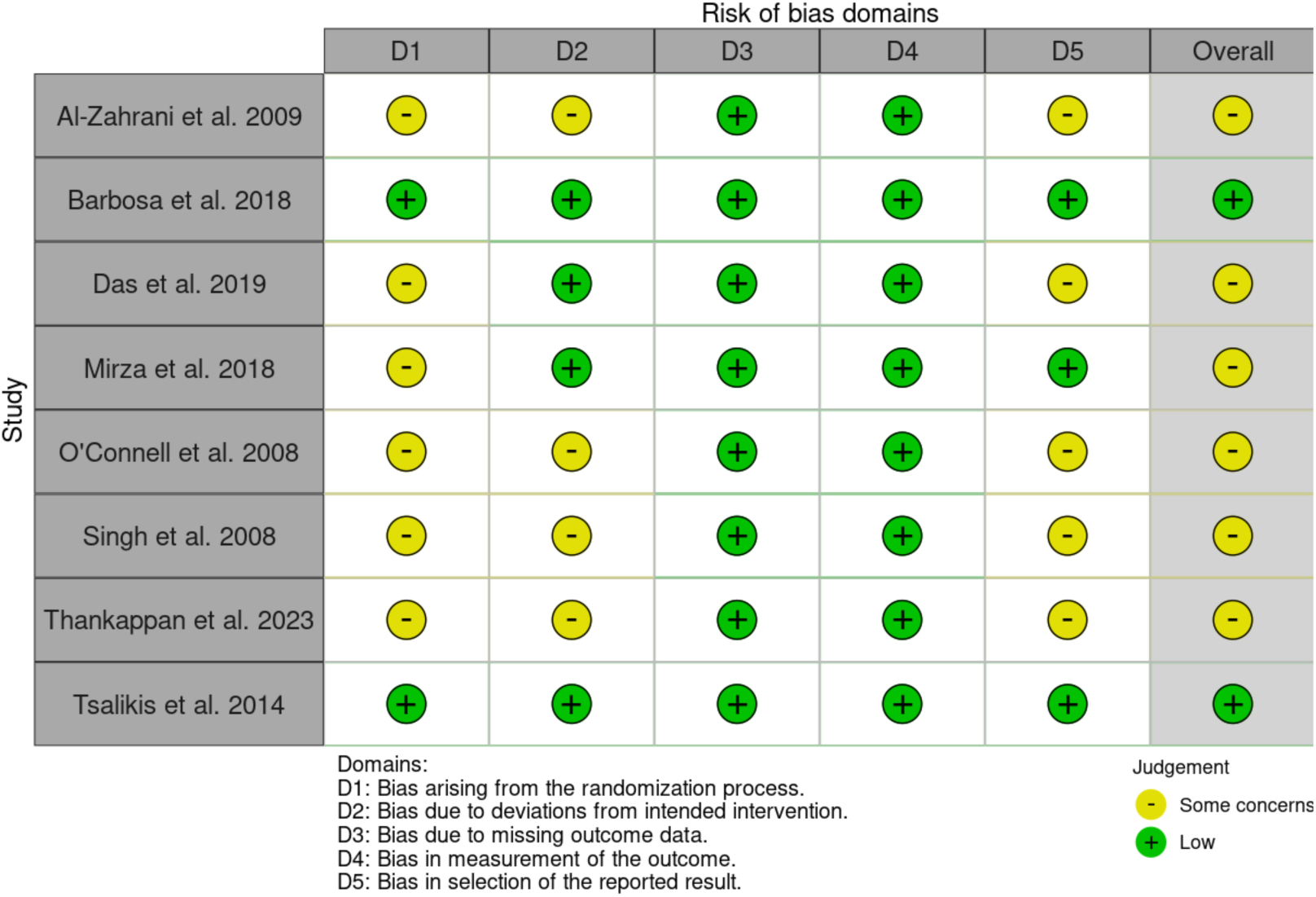
Risk of bias summary of the included studies against five domains.

### Synthesis of Results

For all outcomes assessed in this meta-analysis, four studies with 87 subjects diagnosed with T2DM and periodontitis, assessed the effect of aPDT and SRP compared to SRP alone (Al-Zahrani et al., 2009; Barbosa et al., 2008; Mirza et al., 2018; Thankappan et al., 2023). Additionally, five studies with 191 subjects diagnosed with T2DM and periodontitis, assessed the effect of doxycycline and SRP compared to SRP alone (Al-Zahrani et al., 2009, Das et al., 2019, O’Connell et al., 2008, Singh et al., 2008, Tsalikis et al., 2014).

### Glycated Haemoglobin (HbA1c)

The subgroup combining aPDT with SRP demonstrated a non-significant reduction in HbA1c levels, compared to SRP alone (MD = -0.09 (95% CI: -0.86, 0.68); p = 0.82) (figure 6). In the subgroup where doxycycline was combined with SRP, a non-significant increase in HbA1c was found, compared to SRP alone (MD = 0.02 (95% CI: -0.37, 0.42) p = 0.90) (figure 6). When comparing the effects of these two adjunctive periodontal therapies, the test for subgroup differences did not indicate a statistically significant difference between aPDT and SRP and doxycycline and SRP (p = 0.79). This implies that neither treatment demonstrated a statistically significant advantage over the other in terms of HbA1c reduction. Furthermore, the subgroup combining aPDT with SRP demonstrated significant moderate heterogeneity (I^2^ = 64%; p = 0.04), while the subgroup combining doxycycline with SRP also demonstrated moderate but non-significant heterogeneity (I^2^ = 46%; p = 0.12), as shown in figure 6.

**Figure 6:**
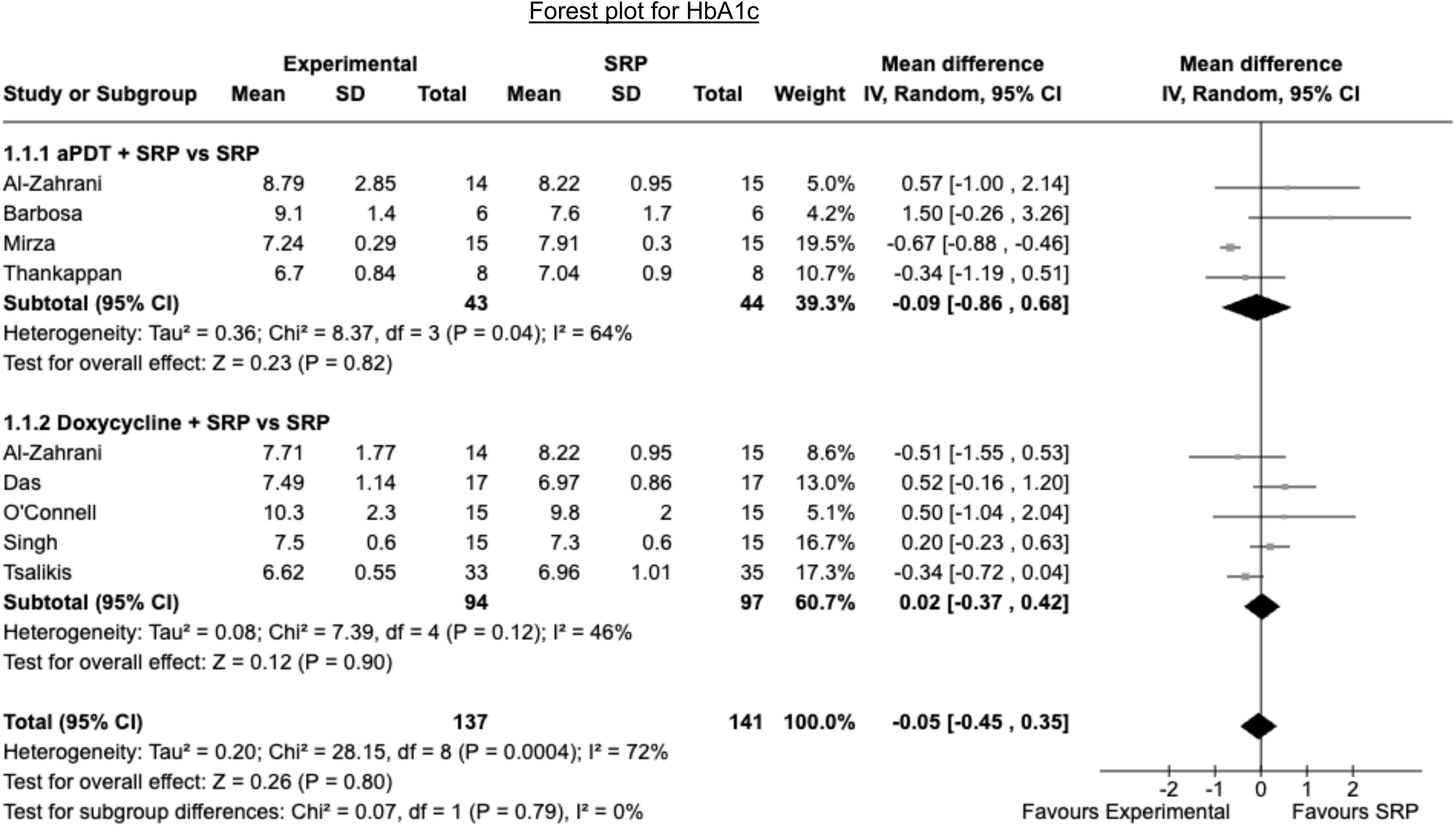
A forest plot comparing the effectiveness of antimicrobial photodynamic therapy (aPDT) versus doxycycline (doxy), both in combination with scaling and root planing (SRP), and compared to SRP alone, on HbA1c levels.

The funnel plot for assessing publication bias, as depicted in figure 7, displayed a degree of asymmetry, with a gap on the bottom left side. There was also a lack of larger studies, comparing aPDT combined with SRP versus SRP, reporting unfavourable outcomes for HbA1c. This was further investigated using statistical tests for publication bias, with the Egger’s regression test for the aPDT and SRP vs SRP group indicating significant asymmetry (p= 0.006), and the fail-safe N suggesting that only a few additional studies could impact the results of the meta-analysis (N = 4; p = 0.011). Additionally, the trim and fill method identified two potentially missing studies. In contrast, the subgroup assessing doxycycline and SRP vs SRP did not show evidence of publication bias when statistical tests for publication bias were employed.

**Figure 7:**
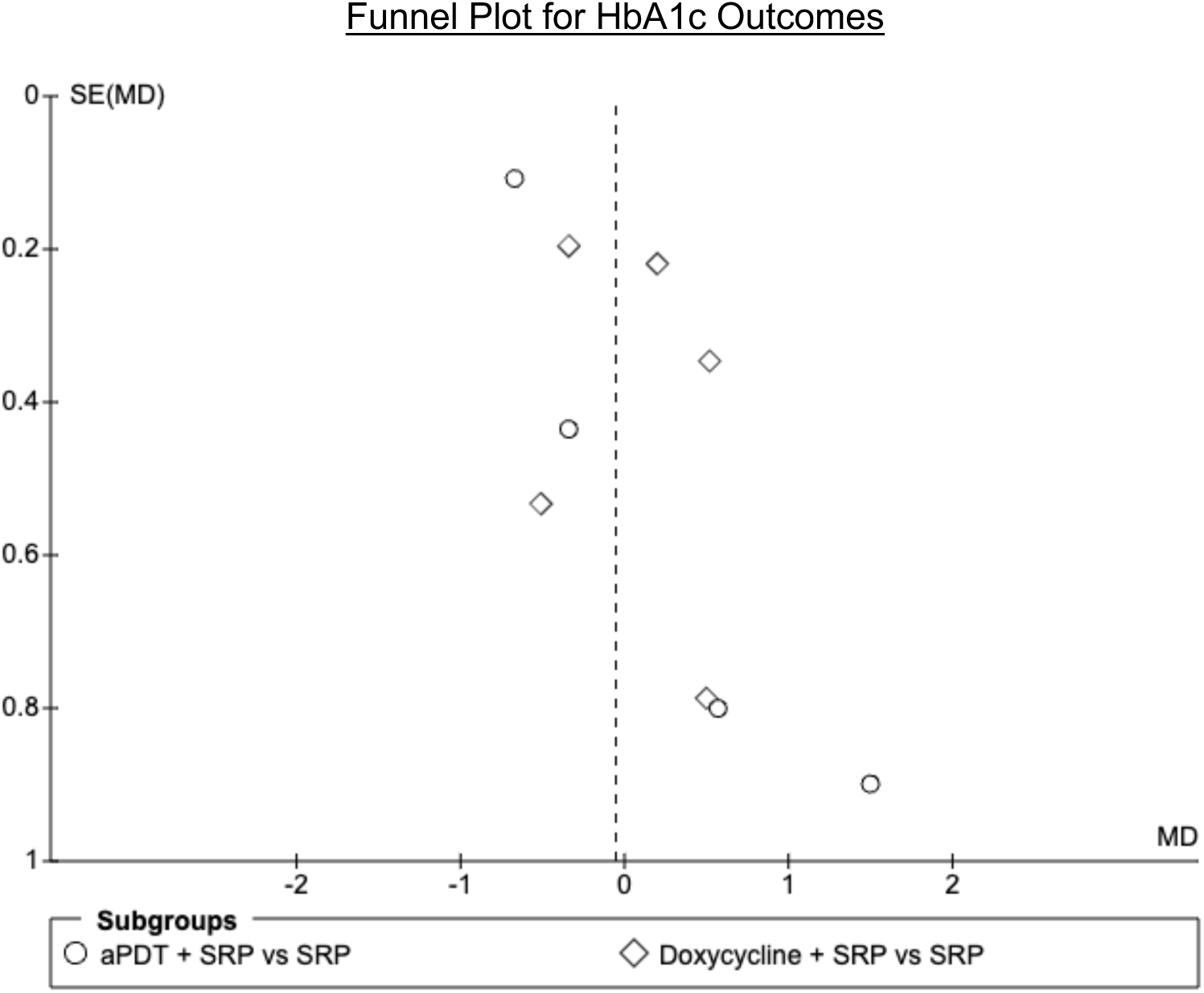
A funnel plot for publication bias in the measurement of HbA1c with the mean difference (MD) versus the standard error of the MD SE(MD) for each study. The vertical line indicates the pooled effect. Circles show antimicrobial photodynamic therapy (aPDT) with scaling and root planing (SRP), and diamonds represent doxycycline with SRP.

### Probing depth

The subgroup combining aPDT with SRP had a statistically significant reduction on probing depth (MD = -0.55 (95% CI: -1.03, -0.07); p = 0.02), as illustrated in the forest plot in figure 8. Conversely, the subgroup combining doxycycline with SRP had a minor, non-significant reduction on probing depth (MD = -0.03 (95% CI: -0.19, 0.13); p = 0.71). The test for subgroup differences revealed a significant difference between the two subgroups (p = 0.04). This implied that through the indirect evidence, aPDT combined with SRP was more effective at reducing probing depth compared to doxycycline combined with SRP. However, the subgroup combining aPDT with SRP demonstrated significant moderate heterogeneity (I2 = 61%; p = 0.05), while the subgroup combining doxycycline with SRP also demonstrated moderate but non-significant heterogeneity (I2 = 43%; p = 0.13).

**Figure 8:**
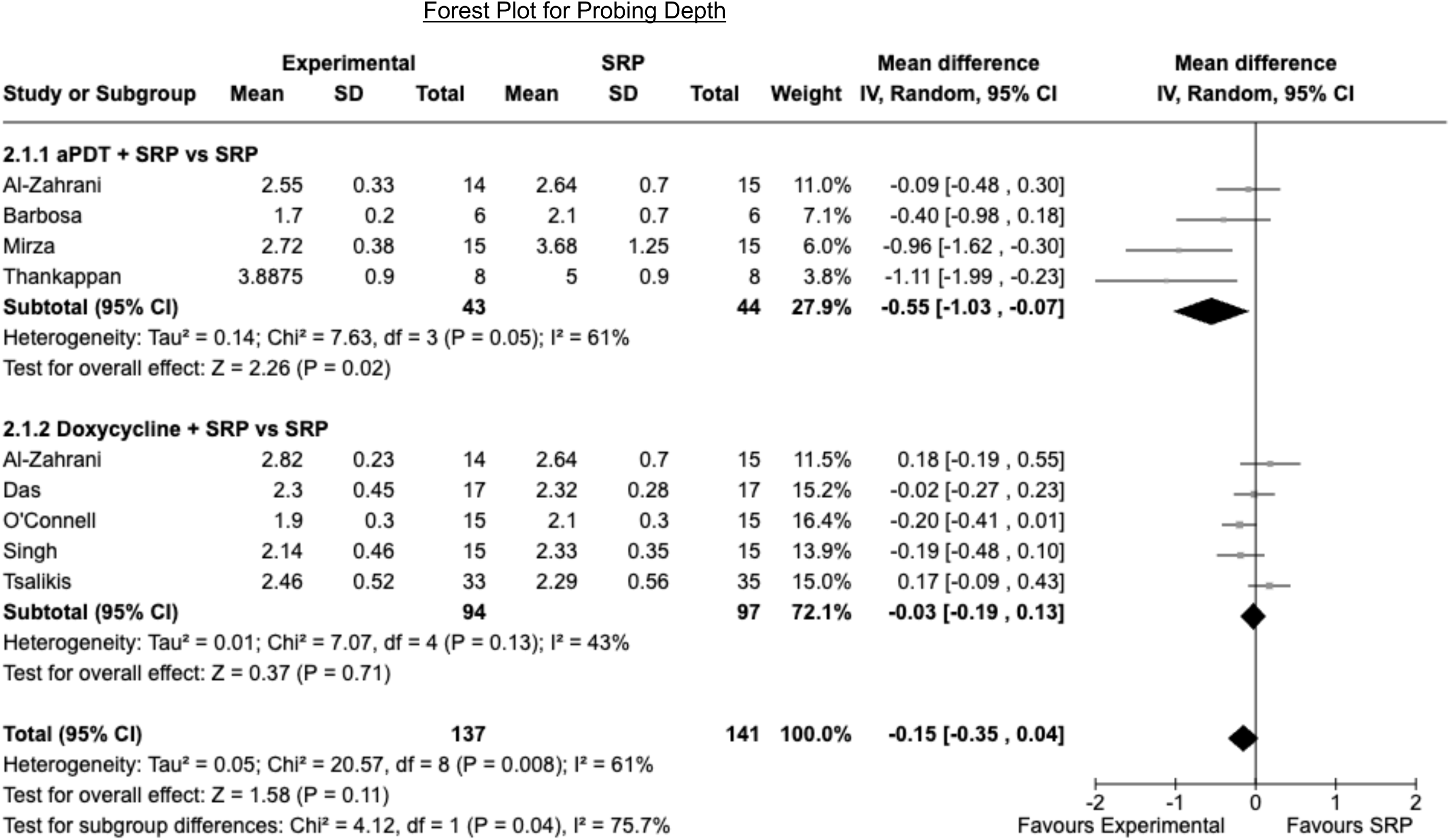
A forest plot comparing the effectiveness of antimicrobial photodynamic therapy (aPDT) versus doxycycline (doxy), both in combination with scaling and root planing (SRP), compared to SRP alone, on probing depth.

The funnel plot for the studies assessing probing depth, shown in figure 9, illustrated considerable asymmetry. There was a lack of smaller studies on the bottom right of the graph for the aPDT and SRP vs SRP subgroup. Statistical tests revealed that the aPDT and SRP vs SRP group had a potential publication bias (fail-safe N = 15 (p<0.01); Egger’s regression (p = 0.009)), with the trim and fill method suggesting one missing study. On the other hand, the statistical tests for publication bias in the doxycycline and SRP vs SRP subgroup, revealed no significant evidence of publication bias.

**Figure 9:**
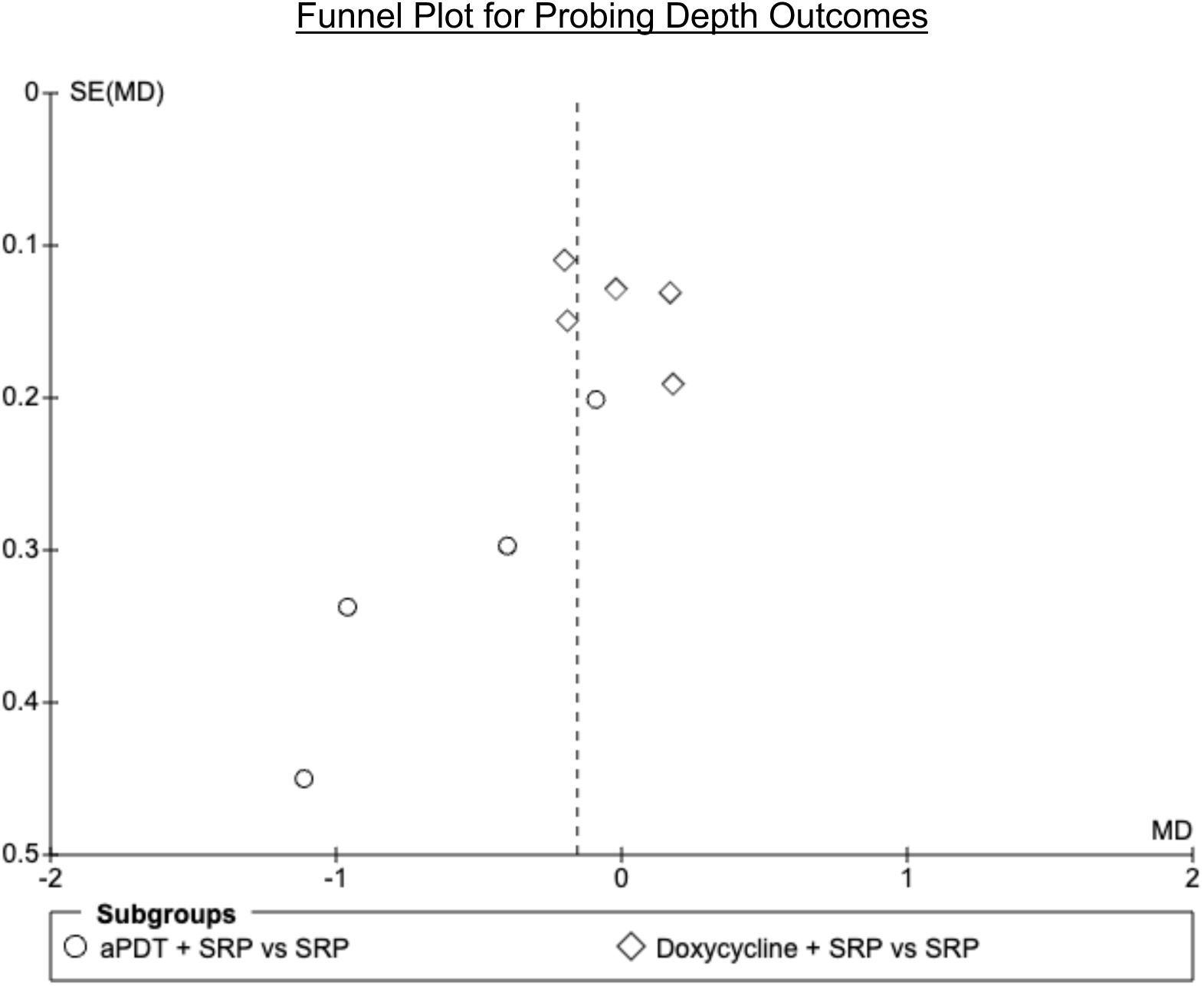
A funnel plot for publication bias in the measurement of probing depth with the mean difference (MD) versus the standard error of the MD SE(MD) for each study. The vertical line indicates the pooled effect. Circles show antimicrobial photodynamic therapy (aPDT) with scaling and root planing (SRP), and diamonds represent doxycycline with SRP.

### Clinical attachment level

Compared to SRP alone, aPDT combined with SRP (MD = -041, 95% CI: -0.99, 0.16) and doxycycline combined with SRP (MD = -0.21, 95% CI: -0.42, -0.01) were found to have a reduction on the clinical attachment level, as shown in figure 10. However, the effect of aPDT combined with SRP compared to SRP alone was non-significant (p = 0.16). Conversely, the effect of doxycycline combined with SRP compared to SRP alone was statistically significant (p = 0.04). Yet, when testing for subgroup differences, no significant differences between the two treatments were found (p = 0.52). This suggests that through indirect evidence, neither subgroup demonstrated a statistically significant advantage over the other in terms of a reduction in clinical attachment level. Moreover, the subgroup combining aPDT with SRP demonstrated a moderate, but non-significant heterogeneity (I^2^ = 52%; p = 0.10), while the subgroup combining doxycycline with SRP demonstrated no heterogeneity (I^2^ = 0%).

**Figure 10:**
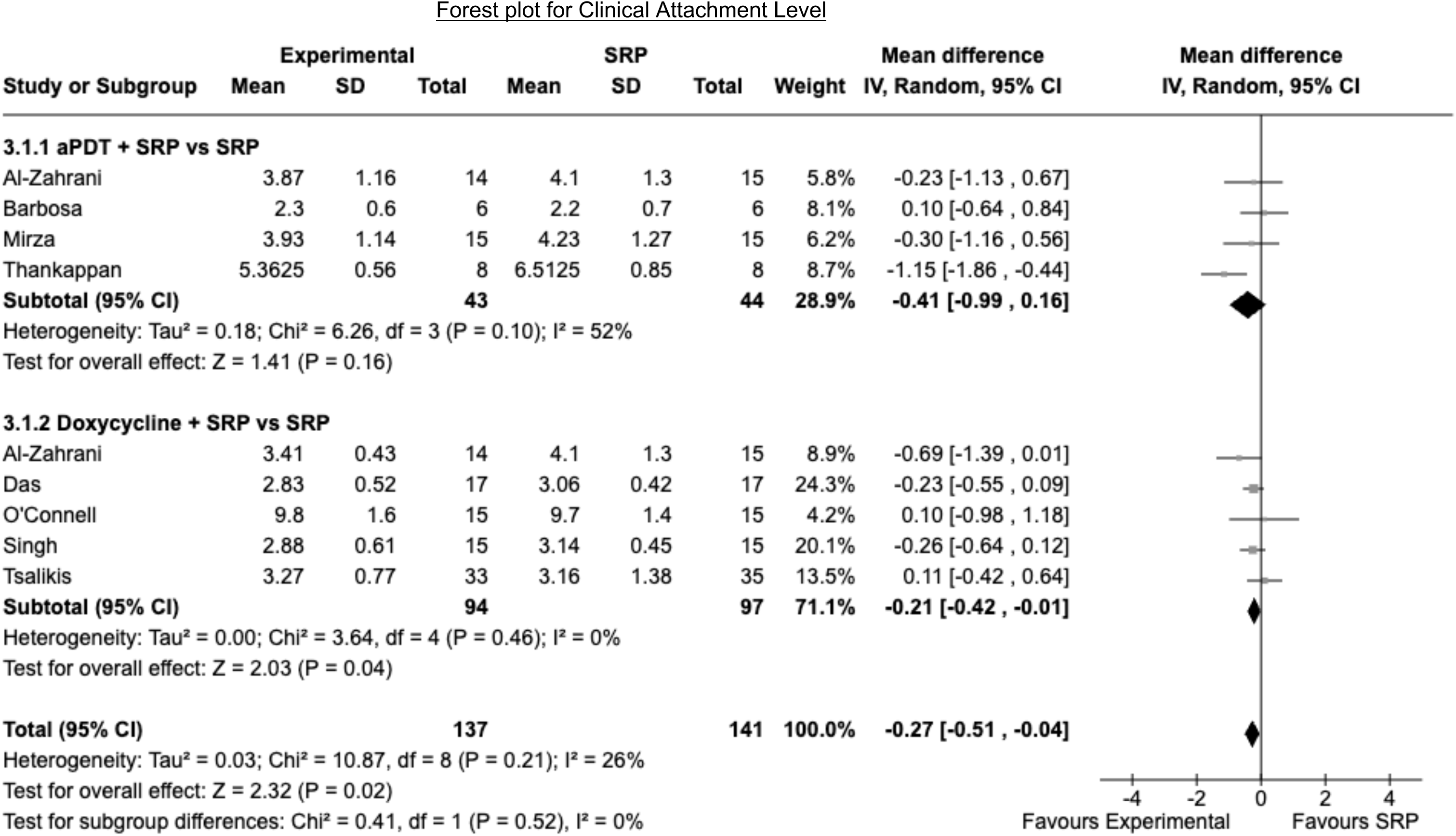
A forest plot comparing the effectiveness of antimicrobial photodynamic therapy (aPDT) versus doxycycline (doxy), both in combination with scaling and root planing (SRP), and compared to SRP alone, on clinical attachment level.

The funnel plot shown in figure 11, for the studies assessing clinical attachment level, revealed a lack of studies, with larger standard errors, for both groups. Statistical tests for publication bias revealed that the aPDT and SRP vs SRP subgroup had a potential concern for bias, though this was not conclusive. The fail-safe N suggested that a small number of additional null effect studies could influence the outcome of the meta-analysis (fail-safe N = 3; p = 0.02). However, the Egger’s regression test showed non-significant results, with the trim and fill method identifying one potentially missing study, indicating a minor bias. Likewise, the fail-safe test for the doxycycline and SRP vs SRP group suggested a slight sensitivity to the inclusion of additional studies (fail-safe N = 2; p = 0.03), though the Egger’s regression test was non-significant, and the trim and fill method did not identify any missing studies. This suggests that there was an absence of significant publication bias in this subgroup.

**Figure 11:**
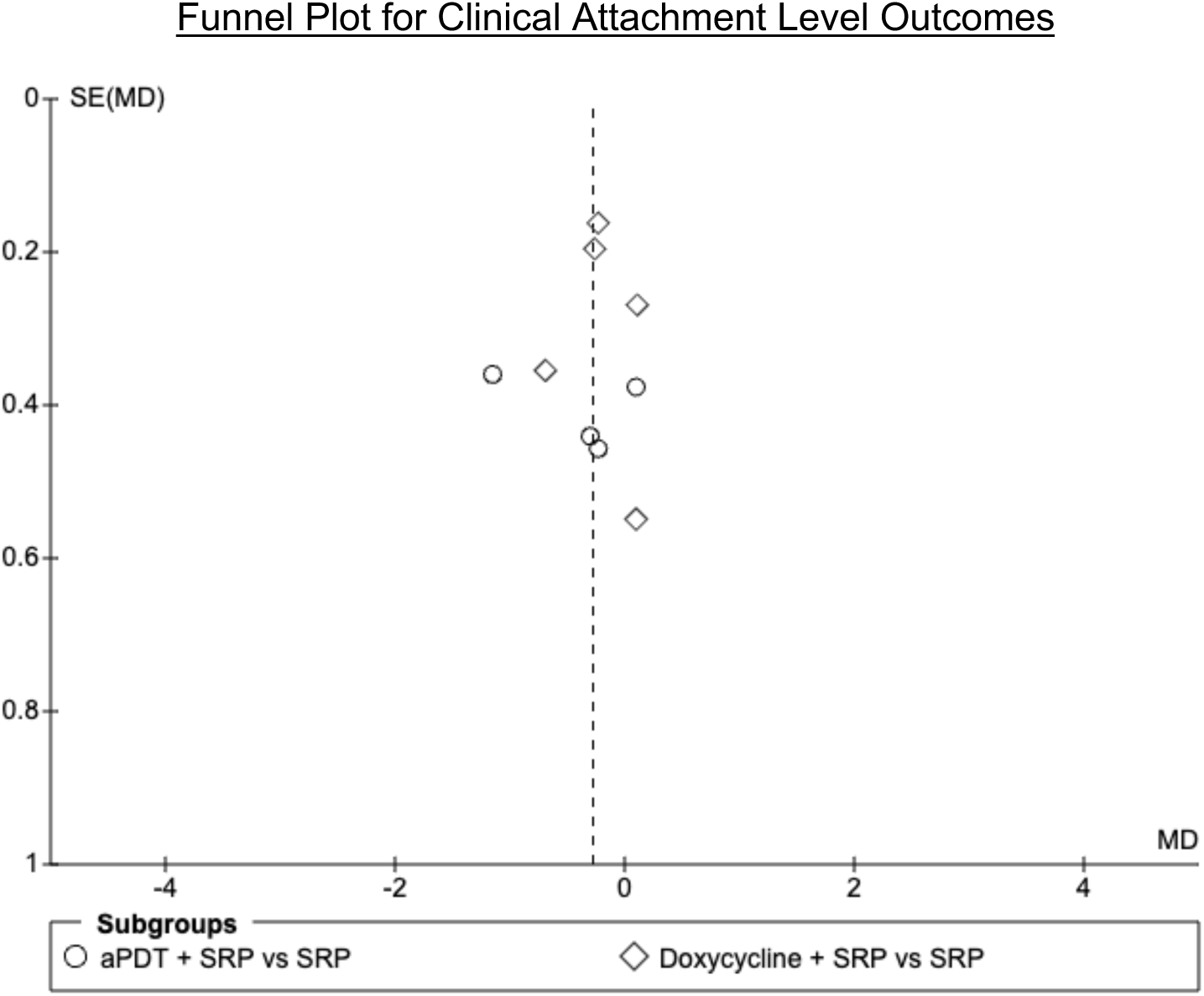
A funnel plot for publication bias in the measurement of clinical attachment level with the mean difference (MD) versus the standard error of the MD SE(MD) for each study. The vertical line indicates the pooled effect. Circles show antimicrobial photodynamic therapy (aPDT) with scaling and root planing (SRP), and diamonds represent doxycycline with SRP.

## Discussion

### Overview of Key Findings

The results of this meta-analysis supported the hypothesis that aPDT could match or outperform doxycycline in improving glycaemic control and periodontal health. In terms of probing depth reduction, aPDT outperformed doxycycline, as evidenced by the significant reduction in the aPDT group compared to doxycycline. Conversely, neither intervention significantly differed from SRP alone or from each other in improving glycaemic control, as indicated by HbA1c levels measured three months post-treatment. Although neither treatment was effective in improving glycaemic control, the results still support the hypothesis, as no significant differences were observed between the two interventions, indicating that aPDT performed similarly to doxycycline in this regard.

However, the effects on clinical attachment level were more nuanced. While aPDT did not show statistically significant improvements compared to SRP alone, likely due to wider confidence intervals reflecting variable treatment effects, doxycycline demonstrated a significant reduction in clinical attachment level compared to SRP alone, with narrower confidence intervals. This suggests that while aPDT may match doxycycline in some respects, such as probing depth and glycaemic control, it may not be as effective in improving clinical attachment level. The variability in aPDT results may be due to unpredictable responses in patients with uncontrolled T2DM. Overall, these findings partially support the hypothesis, particularly regarding probing depth reduction, but also highlight the limitations of aPDT in other measures like clinical attachment level.

In comparison to the wider literature, several meta-analyses have explored the effect of adjunctive periodontal therapies for T2DM patients. Like this meta-analysis, Corbella et al. 2022 found that aPDT, compared to SRP alone, enhanced probing depth reductions and had no significant effect on clinical attachment levels. However, the results from Corbella et al. 2022 also diverged from this meta-analysis, reporting improvements in HbA1c. This is possibly due to their inclusion of a range of aPDT methods leading to differences in bacterial reduction compared to this meta-analysis. Additionally, other meta-analyses have compared the effects of aPDT with SRP alone, but contradict these findings, observing no significant differences between the two groups (Al-Hamoudi, 2017; Abduljabbar et al., 2017). Additionally, studies have examined the effects of antibiotics compared to SRP alone (Yap & Pulikkotil, 2019; Grellman et al., 2016), finding mixed outcomes and contrasting with the significant effect observed on clinical attachment level in this meta-analysis. The differences between published meta-analyses and this meta-analysis may stem from the inclusion and exclusion criteria. For example, the Al-hamoudi. 2017 meta-analysis included T2DM patients who were also cigarette smokers, while this meta-analysis excluded cigarette smokers due its risk factor on periodontal outcomes (Bunaes et al., 2015; Guo & Di-Pietro, 2020).

The study by Ramos et al. 2015 stands out as the only RCT to date that directly compares aPDT and doxycycline as adjuncts to SRP in T2DM patients. However, its findings diverge from this meta-analysis regarding glycaemic control improvements. While the RCT reported significant HbA1c improvements in both treatments, this meta-analysis found no significant changes. This discrepancy may arise from the different study designs: Ramos et al. 2015 conducted a head-to-head RCT, while this meta-analysis used indirect comparisons, potentially offering a broader but less direct perspective. Nonetheless, both the RCT and this meta-analysis agree that aPDT as an adjunct to SRP was more effective in improving probing depth, reinforcing its potential benefit for treating T2DM patients with periodontitis.

### Mechanisms Behind the Findings

The superior performance of aPDT in reducing probing depth, could be attributed to several factors. While both adjunctive treatments target periodontitis, aPDT focuses on reducing microbial reduction within periodontal pockets and controlling local inflammation, while doxycycline inhibits matrix metalloproteinases (MMPs) (Srinath, 2015) and exerts systemic anti-microbial effects (Prakasam et al., 2012). Being a local therapy, aPDT may penetrate deeper into the periodontal tissues, effectively targeting pathogens and reducing inflammation within the periodontal pockets. On the other hand, doxycycline is a systemic therapy, which may have limited penetration into periodontal tissues. This could be a potential explanation for the reduced efficacy of doxycycline in reducing probing depth observed in this meta-analysis. Furthermore, by reducing the microbial load, aPDT may also lower the levels of local pro-inflammatory cytokines within periodontal pockets (Vivas et al., 2016), which could potentially reduce systemic inflammatory markers that affect insulin resistance (Kiely et al., 2007) and indirectly influence HbA1c levels. However, as previously noted in this meta-analysis, a reduction in HbA1c was not found. The literature often considers a three-month timeframe as short-term in the context of periodontal therapy (Ramos et al., 2015; Yap & Pulikkotil, 2019; Navarro-Sanchez et al., 2007). Given this perspective, it could be suggested that the studies included in this meta-analysis, all with three-month timelines, might not have been sufficiently long enough to allow for significant improvements in glycaemic control to manifest.

### Limitations of this Meta-analysis

Heterogeneity was found in the subgroup comparing aPDT and SRP with SRP alone for HbA1c and probing depth, while the subgroup comparing doxycycline and SRP with SRP alone did not show any significant heterogeneity. This suggests that the effect of aPDT was not consistent across the studies. However, given the unpredictable nature of T2DM patients’ responses to periodontal therapy (Geraldo et al. 2023; Lalla et al., 2000; Graves et al., 2007), it is plausible that this may have caused true variations in treatment effects, contributing to the observed heterogeneity. Furthermore, publication bias was detected, particularly in the subgroup comparing aPDT and SRP with SRP alone. This bias may have stemmed from selective reporting of studies with positive outcomes or the underrepresentation of studies reporting unfavourable results. However, this could also be because aPDT is a relatively new approach in periodontal treatment (Takasaki et al., 2009) and its use has not yet been recommended for periodontal therapy in practices, due to a lack of scientific evidence (Sanz et al., 2020). Therefore, there may be fewer studies available, which could contribute to a lack of data and the observed publication bias. In addition, the specific focus of T2DM patients might have further restricted the number of available studies for inclusion in this meta-analysis.

A key limitation of this meta-analysis was the use of post-treatment values instead of changes from baseline within each group. This approach might underestimate the effectiveness of treatments, as seen in included studies (Singh et al., 2008; O’Connell et al., 2008), where doxycycline groups showed greater HbA1c reductions compared to SRP alone. However, these results appeared less significant in this meta-analysis due to higher baseline HbA1c levels in doxycycline groups. This meta-analysis also faced limitations due to its reliance on indirect comparisons, where slight variations in control groups (SRP alone) across studies could have diluted the power of original randomisation. Since these studies were not designed to be compared with each other, this may introduce bias and heterogeneity (Bucher et al., 1997). Therefore, these findings should be interpreted with caution.

### Clinical Implications

The significant reduction in probing depth observed with aPDT emphasises its clinical relevance in improving periodontal health, suggesting that tailored treatment approaches may be beneficial in T2DM patients. Given the variable responses of T2DM to standard periodontal therapies (Geraldo et al. 2023; Lalla et al., 2000; Graves et al., 2007), incorporating adjunctive therapies like aPDT, could improve periodontal outcomes. Based on the results of this meta-analysis, clinicians should prioritise the continued use of diabetes medications to maintain glycaemic control in patients with T2DM and not solely rely on periodontal therapy as an alternative method for glycaemic control. An integrated treatment strategy that includes diabetes medications, such as metformin, and aPDT as an adjunct to SRP could offer an effective alternative to antibiotics in managing periodontitis in T2DM patients. This approach could not only aid in diabetes management, but also help reduce the risk of antibiotic-resistant bacterial infections, addressing a critical global health concern (WHO, 2020).

### Recommendations for Future Research

Given that this meta-analysis found that doxycycline was ineffective in two out of three outcomes when used as an adjunctive to SRP, it is necessary to conduct a thorough re-evaluation of its use in periodontal treatment. Future research could focus on confirming these findings, exploring whether specific patient groups might benefit from this adjunctive treatment, and assess the potential risk of antibiotic resistance. Following the re-evaluation of doxycycline, more research could advance to comparing doxycycline to aPDT using direct evidence from RCTs, with further meta-analyses pooling these results to provide a clear comparison and addressing the potential biases of this current meta-analysis. These studies should be conducted for at least six months to assess the long-term effects of these treatments, especially regarding glycaemic control. By addressing these areas of research, the field could find more effective treatment options for periodontitis in the context of T2DM patients.

## CONCLUSION

This meta-analysis assessed the efficacy of antimicrobial photodynamic therapy (aPDT) versus doxycycline, both alongside scaling and root planing (SRP) for treating periodontitis and managing glycaemic control in patients with T2DM. The findings from this meta-analysis revealed that aPDT, when used as an adjunct to SRP, effectively reduces probing depth, a parameter reflective of periodontitis. This highlights aPDT’s potential as a non-antibiotic option for enhancing periodontal treatment outcomes in T2DM patients. Conversely, the variable effects associated with doxycycline, combined with concerns regarding antibiotic resistance, suggests that its use should be re-evaluated and applied with caution. Moreover, the findings of this study suggest that relying solely on short-term periodontal therapy for improving glycaemic control is insufficient. In conclusion, this meta-analysis indicates that aPDT can match and exceed the efficacy of doxycycline for treating periodontitis in patients with T2DM. However, achieving optimal glycaemic outcomes in T2DM patients with periodontitis may require an integrated approach that includes periodontitis treatment alongside diabetes medications.

## Data Availability

The data used in the meta analysis were extracted from various electronic databases including PubMed Scopus CENTRAL Embase and MEDLINE.

https://doi.org/10.1902/jop.2009.090206

https://doi.org/10.1016/j.pdpdt.2018.04.013

https://doi.org/10.1016/j.pdpdt.2019.08.003

https://doi.org/10.4103/jisp.jisp_7_23

https://doi.org/10.5005/jp-journals-10024-2722

https://doi.org/10.1902/jop.2008.070250

https://doi.org/10.1111/jcpe.12287

https://doi.org/10.4103/0973-3930.43097

